# Direct measurement of motor unit twitch properties using routine clinical ultrasound and optical flow

**DOI:** 10.64898/2026.10.26.26366517

**Authors:** Stuart Maitland, James Scott, Karen Suetterlin, Roger G Whittaker

## Abstract

**Introduction/Aims:** Current motor unit assessment techniques overlook the mechanical output that underpins function and are limited: surface recordings are weighted to the superficial ∼1cm and needle EMG to a ∼2 mm radius. We investigated whether clinical muscle ultrasound can measure motor unit twitches directly.

**Methods:** Peripheral nerve stimulation targeted abductor digiti minimi (ADM), abductor pollicis brevis (APB) and tibialis anterior (TA). Stimulus intensity was set 10% above supramaximal and decreased in 0.01-0.05 mA steps until the Compound Muscle Action Potential (CMAP) was abolished. An 18 MHz transducer was placed transverse to the muscle, co-located with the active electrode, with video time-locked to each stimulus. Inter-frame motion was quantified using Farneback optical flow, and twitch/CMAP amplitudes compared to derive mechanical and electrical activation thresholds and a motor unit twitch index.

**Results:** We obtained 66 recordings from 22 participants (11 female, 21-63 years); 49 met prespecified quality criteria. Threshold medians (CMAP versus ultrasound) were 6.84/5.82 mA in ADM, 5.28/4.84 mA in APB and 10.34/9.83 mA in TA. The ultrasound threshold was lower than the CMAP threshold in all three muscles (ADM +0.27 mA, p = 0.009; APB +0.08 mA, p = 0.001; TA +0.21 mA, p = 0.011). A motor unit twitch index was measurable in 32/49 recordings: 14.4 in ADM, 14.9 in APB and 18.6 in TA. Median contraction/half-relaxation times for ADM, APB & TA were 80/80, 88/95 and 79/103 ms respectively; trial-to-trial variability was 5.3/8.3 ms.

**Discussion:** Clinical ultrasound can be used to directly measure clinically relevant twitch parameters such as twitch amplitude which recruits in parallel with CMAP and is not detectable with conventional clinical neurophysiology techniques. Mechanical activation is detected at slightly lower stimulus currents than the CMAP, and the twitch signal plateaus before the CMAP does; neither effect was confined to the larger muscle.

## Introduction

Electromyography (EMG) remains the clinical gold standard for assessing motor unit (MU) properties; however, it is fundamentally limited to capturing electrical potentials rather than the resultant mechanical twitch of muscle tissue. Furthermore, EMG modalities face significant spatial constraints. Surface EMG (sEMG) is non-invasive but heavily biased toward the superficial 1 cm of tissue^1^. Conversely, needle EMG (nEMG) offers high spatial resolution but records from a highly restricted radius of approximately 2 mm^2^. Neither approach can directly assess the mechanical twitch of the muscle tissue itself, whose time course (contraction and relaxation) reflects motor unit type and metabolic properties of the muscle fibre population^3,4^.

To capture the mechanical output of motor units, traditional approaches measure limb or joint movement via force transducers or accelerometers. These methods provide an indirect assessment that is heavily confounded by the biomechanics of the joint and tendon^5,6^. Variables such as tendon compliance and joint resistance introduce nonlinearities into the recorded response. Furthermore, these measurements can be skewed by non-neurological factors, including prior musculoskeletal injury^7^, or connective tissue disorders^8^, thereby dissociating the measured external force from the true internal muscle fascicle contraction. Mechanical measurement is not new to clinical neurophysiology. Jolly’s 1895 demonstration of myasthenic fatigability was an observation of the fading mechanical contraction under repetitive stimulation^9^, and Desmedt showed that in Duchenne muscular dystrophy abnormalities in twitch properties, measured with a force transducer, preceded any recordable EMG abnormality in the same muscle^10^. Despite this, mechanical measurement did not enter routine practice. Force transducer recording requires rigid fixation and a purpose-built rig for each muscle, is limited to the few muscles acting on an accessible lever and reports only whole-muscle output; twitch measurement remains a research technique. Over the same period the electrical signal became standardised on commercial equipment and gained motor unit-level resolution through needle EMG and motor unit number estimation techniques. Diagnostic neurophysiology therefore retained Jolly’s protocol but replaced his mechanical readout with an electrical one and the mechanical output of motor units is now essentially ignored in clinical practice.

Recently, muscle ultrasound has emerged as a direct, non-invasive technique for observing mechanical contractile properties in vivo. Examples include utilising high-frame-rate ultrasound in conjunction with spike-triggered averaging^11,12^ or fascicle length tracking during muscle stimulation^13^. While these ultrafast techniques provide excellent temporal resolution for isolating individual twitches, they require specialised, computationally heavy equipment that is currently restricted to research environments.

Standard diagnostic ultrasound is rapidly entering routine clinical neurophysiology practice, but these clinical systems operate at significantly lower frame rates (<150 Frames Per Second (FPS)). We have previously combined clinical ultrasound with nEMG to perform a targeted “Scanning EMG” transect of a single motor unit^14^; however this technique assessed single motor units and did not attempt to measure the entire motor unit population. More recently, others have used ultrasound for measuring fascicular twitch properties^15^, however these have not been yet applied to measuring motor unit populations.

This study aimed to demonstrate that routine clinical ultrasound, when coupled with optical flow analysis, can determine motor unit mechanical activation thresholds and extract mechanical contractile properties.

## Methods

### Participants and ethics

Ethical approval was obtained from the Newcastle University Faculty of Medical Sciences Ethics Committee (Ref: 61737/2023). Written informed consent was obtained from all participants prior to data collection. Healthy volunteers were recruited from a local participant database. Exclusion criteria were a history of diabetes, carpal tunnel syndrome, neurological disease, current pregnancy, or a cardiac pacemaker.

### Instrumentation and data acquisition

Recordings were obtained from three muscles: abductor digiti minimi (ADM), abductor pollicis brevis (APB) and tibialis anterior (TA). Electrical stimulation was delivered as a 0.2 ms square-wave pulse using a Digitimer DS5 constant-current stimulator at 0.7 Hz (Digitimer, Welwyn Garden City, UK). Surface electromyography (EMG) was amplified (gain 1000) with a Digitimer D440 amplifier and sampled at 10 kHz. A CED Micro1401 acquisition unit (Cambridge Electronic Design, Cambridge, UK) served as sequencer for stimulation timing and recording.

Stimulating electrodes (Covidien ECG electrodes, Medtronic, MN) were applied over the relevant peripheral nerve after skin preparation. Surface EMG was recorded with the active electrode one-third along the muscle belly from the proximal end and the reference electrode over the distal tendon.

Ultrasound imaging used a TeleMed ArtUS device (Telemed, Vilnius, Lithuania) with an L18-7H30-A5 linear transducer (18 MHz, 30 mm field width). Imaging depth was set to 20(APB, ADM) or 30 (TA) mm to include the deepest extent of the muscle. The probe was positioned transverse to the muscle fibres, adjacent to the active surface EMG electrode, and held manually with minimal pressure, sufficient for acoustic contact without tissue compression (Figure 1A). For hand muscles, the ultrasound device was held manually by a trained operator, whilst for tibialis anterior a custom-built ball-and-socket strap attachment held the probe to the surface of the skin whilst maintaining position on the skin surface. Gain, focus and dynamic range were optimised for muscle visualisation and held constant within a given muscle.

**Figure 1.**
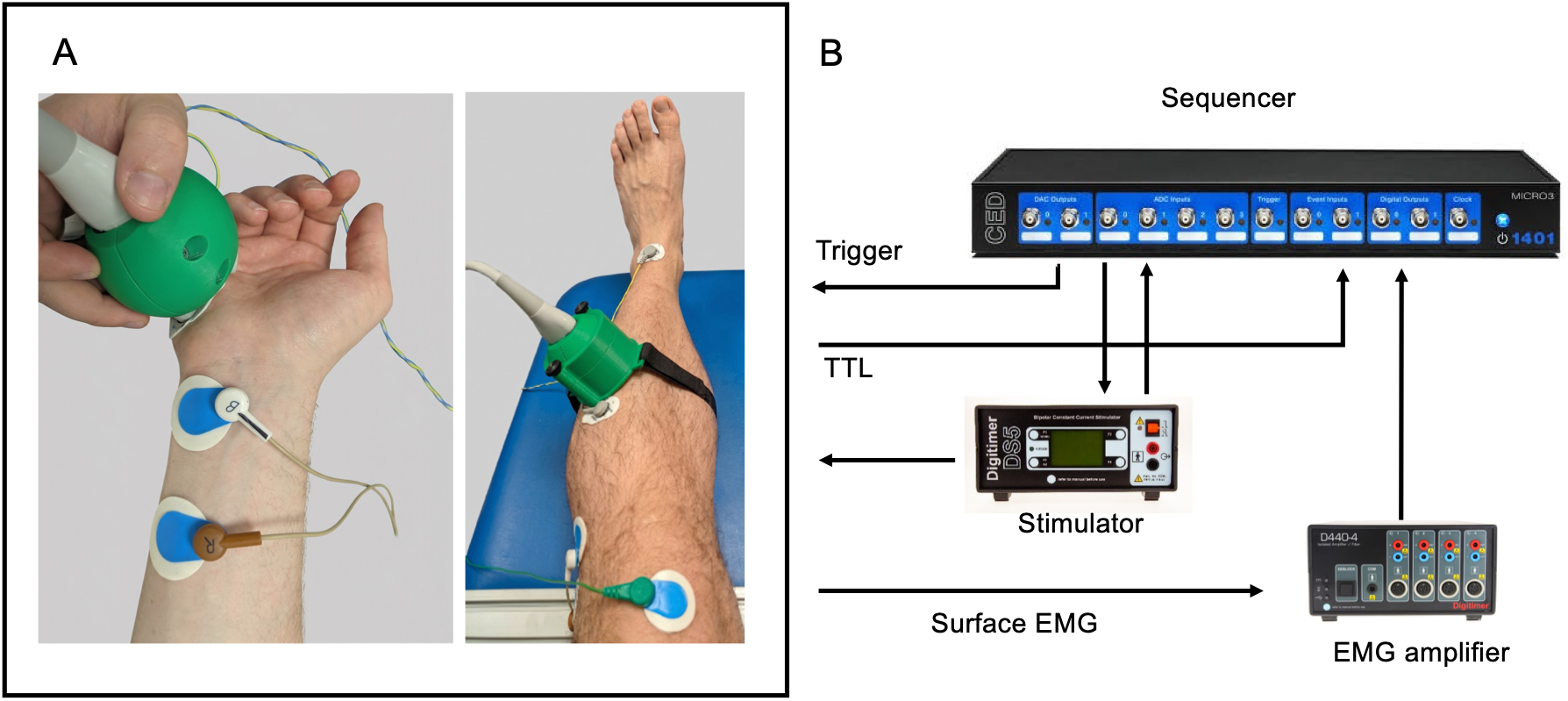
Experimental setup. Panel A-Ultrasound transducer is placed on muscle belly, transverse to muscle fibres, adjacent to active electrode, with reference electrode on distal tendon. Peripheral nerve stimulation is delivered to fibular (tibialis anterior, right), ulnar (abductor digiti minimi, left) or median (abductor pollicis brevis, not shown) nerves. A CED Micro1401 sequencer triggers ultrasound capture and peripheral nerve stimulation via a Digitimer DS5 constant current stimulator, and samples amplified EMG data via a Digitimer D440 and a TTL frame acquisition signal from the ultrasound scanner. TTL = transistor transistor logic pulse.

Ultrasound capture was triggered by a transistor-transistor logic (TTL) pulse from the sequencer, after which frames were acquired at the maximum available rate. The device emitted a falling edge with each frame, recorded on a separate channel alongside sEMG and the stimulus pulse, giving frame-level temporal alignment between video and electrophysiology (Figure 1B). Ultrasound capture was triggered 200ms before electrical stimulation to derive a baseline measurement.

#### Stimulation protocol

Stimulus current was first increased until a supramaximal compound muscle action potential (CMAP) was obtained, then set 10% above that level. A decrementing series of stimuli was delivered in steps of 0.01–0.05 mA with a hold of 3 stimuli per level, the step size chosen to obtain at least 50 stimuli across the activation range while limiting participant discomfort.

### Signal processing

Analysis used a custom pipeline in Python 3.13 (numpy 2.2.5, scipy 1.15.3, pandas 2.2.3, OpenCV 4.11.0). EMG was notch-filtered at 50 Hz and band-pass filtered 5–3000 Hz. CMAP amplitude was measured peak-to-baseline in a 2–20 ms post-stimulus window, with baseline taken 150–50 ms before the stimulus.

Video–EMG alignment was verified per recording by comparing the number of measured TTL pulses against the number of video frames; recordings with a mismatch of frames and ultrasound TTL pulses were rejected as having insufficiently certain alignment.

#### Motion quantification

Muscle motion between frames was quantified with the Farneback dense optical flow algorithm^16^ (OpenCV^17^). Flow was projected onto the dominant motion direction rather than taken as unsigned magnitude, preserving contraction-relaxation direction. The resulting trace was high-pass filtered (second-order Butterworth, 1.0 Hz) to remove slow drift arising from probe movement.

Ultrasound produces significant speckle noise, and optical flow measurements will detect these as constant movement. This produces a noise floor to motion quantification, which affects the reliable detection and quantification of true tissue motion. To overcome these limitations, twitch amplitude was measured as a signed spatiotemporal matched-filter projection of the tissue deformation field rather than as a peak of motion velocity.

A canonical spatial flow template was therefore constructed for each recording. Using this approach, spatially incoherent speckle projects to approximately zero mean with much reduced variance, whereas coherent muscle motion adds constructively; the resulting amplitude is signed. Per-trial baseline noise was quantified from the median absolute deviation of the pre-stimulus samples of the projection trace.

Template construction required at least 5 trials exceeding the noise threshold. Where this was not met, no template could be built, and recordings without a validated template were excluded. Chunks with a spatial coherence score below 0.1 were discarded before curve fitting, and trials whose pre-stimulus noise exceeded 5 times the cohort median were flagged as movement artifact.

#### Twitch timing

Motion onset for each twitch was defined as the point after stimulation at which velocity exceeded 10% of peak velocity. Detected peaks were averaged over ±2 frames.

Since the motion is signed, the first positive phase represents contraction, and the negative phase represents relaxation (Figure 2A & 2B). From this, two timing parameters were derived per trial:

**Figure 2.**
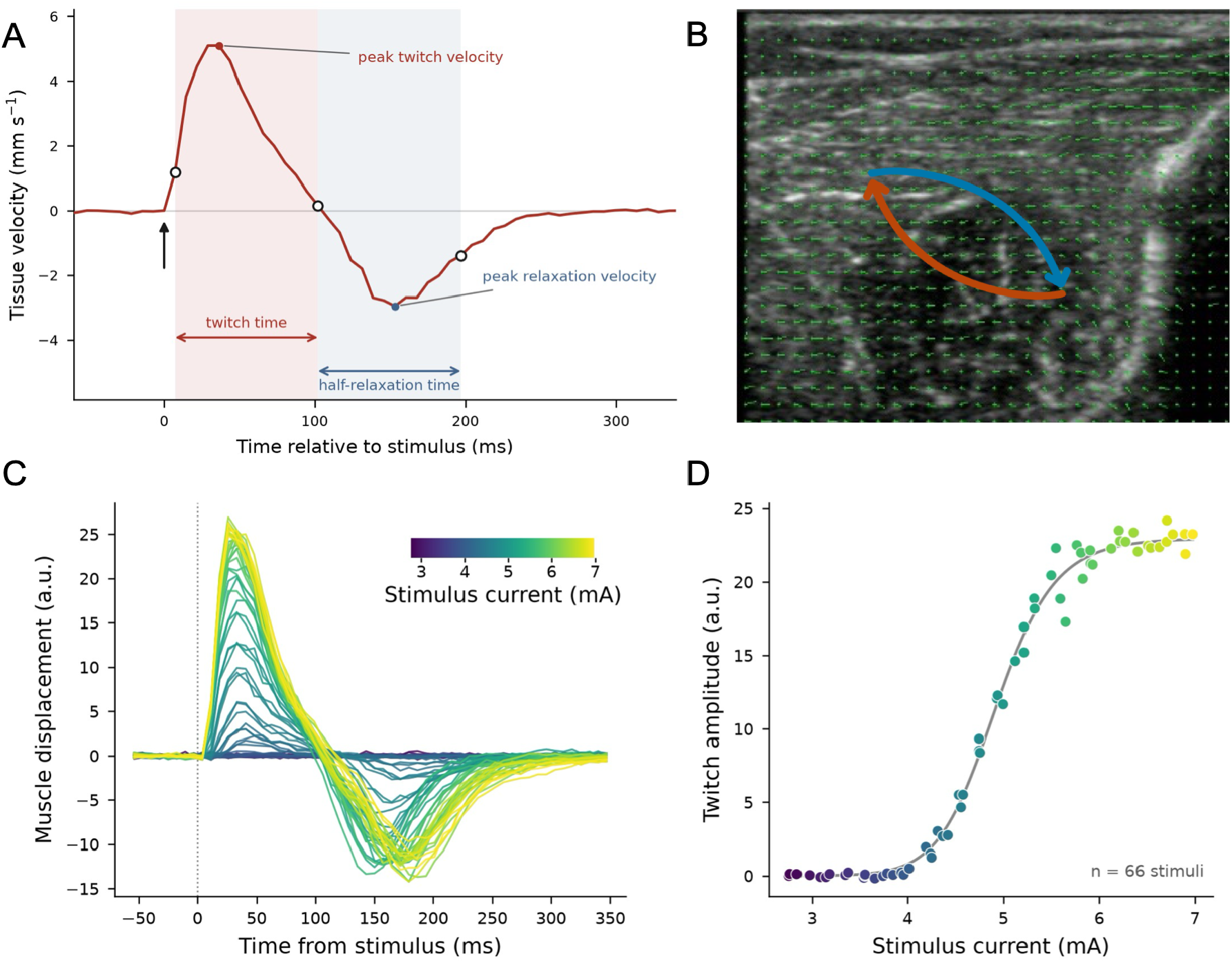
Example twitch profile (A) showing the twitch and relaxation phases of muscle activation, derived from the dominant tissue motion direction (B). The amplitude of the twitch displacement across varying stimuli strengths (C) can be plotted to produce a recruitment curve (D)

– **Contraction time**: onset of motion to the first return to zero, terminating the contraction phase.
– **Half-relaxation time**: from end of contraction to half-recovery toward baseline.

Per-trial values were aggregated within each recording using the median, with outliers removed at 1.5 x IQR.

### Recruitment analysis

#### Primary: model-free thresholds

For each modality, the response was binned by stimulus current. The low-current reference level was taken as the median of the lowest 5% of stimulus bins and the plateau as the 95th percentile of the binned response. The activation threshold was defined as the current at which the response first reached 5% of this reference-to-plateau range.

#### Secondary: parametric (4PL) replication

As a parametric replication, a four-parameter logistic was fitted to each stimulus–response curve (CMAP and twitch) by non-linear least squares(SciPy curve_fit),:

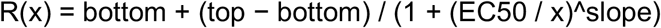

For each recording and modality, trials were grouped by delivered stimulus current and the median response at each current level was taken as the stimulus–response curve. The four-parameter logistic above was fitted to these medians with bottom and top initialised at the minimum and maximum observed response and constrained to lie within that range ± 20 % of its span. EC50 is the current at which the fitted response lies midway between bottom and top, i.e. the midpoint of the recruitment curve.

### Motor unit twitch index

An index of the number of motor units producing muscle twitch was derived from the fitted mechanical recruitment curve. Four monotone models were fitted to every twitch curve; Hill (equivalent to the 4PL above), Gompertz, Weibull and probit. Slope parameters were bounded under a common ceiling on normalised maximum slope, so that differences between models are not artifacts of incomparable bounds. Two derivation rules were applied to each model, and all eight model × rule combinations were computed for every recording.

The primary estimate is Hill / Rule B: index = fitted top / single motor unit twitch (SMUT), where SMUT is the median increment in the sub-EC50 recruiting region exceeding twice the recording’s own measured noise floor (Figure 2C). Bootstrap confidence intervals were computed by 200 stratified resamples. Recordings with fewer than five resolvable increments are reported as unmeasured. The alternative Rule A (top ÷ mean positive increment) is reported alongside.

This estimator has the form of incremental motor unit number estimation^18^: a maximal response divided by a representative single-unit increment sampled from the tail of a recruitment curve. Three properties of the ultrasound mechanical signal mean this should be read as an index rather than a count. Firstly, an increment is accepted only if it exceeds twice the recording’s noise floor, so the smallest twitches do not contribute to the SMUT estimate, and accepted increments may represent more than one unit. Second, as in all incremental methods, near-threshold axons fire probabilistically, so the increment at a given current may comprise different units from trial to trial (alternation). Third, twitch displacement summates non-linearly, whereas CMAP amplitude summates approximately linearly. Values are therefore used for within-cohort comparison and ranking only and are reported with bootstrap intervals and model-to-model spread.

### Quality control and exclusions

1. **Curve stability:** Recordings were rejected if the EC50 coefficient of variation (EC50 standard error ÷ |EC50|) reached 0.35, or if EC50 exceeded the maximum delivered current.
2. **Validated amplitude template:** Recordings without a spatial template (fewer than 5 trials exceeding a coherence score of 0.70) were excluded.
3. **Stimulus-locked motion:** recordings whose mean supramaximal motion trace showed no stimulus-locked peak (peak contrast below 6 SD of the pre-stimulus trace and post-

/pre-stimulus motion ratio below 1) were excluded.

### Statistical analysis

Between-modality comparisons within a muscle were paired within recording and tested with the Wilcoxon signed-rank test, since the distributions are skewed. P-values were adjusted with the Benjamini–Hochberg procedure within the family of six primary within-muscle tests (threshold and plateau in three muscles). Exact p-values are reported throughout. Analysis was performed in Python 3.13 with scipy 1.15.3.

## Results

### Cohort

66 recordings were obtained from 22 participants (n=11 female). The mean age of participants was 28, SD 9, range 21-63. 17 recordings did not meet the prespecified quality-control criteria: 13 because fewer than five trials reached the spatial coherence threshold required for template construction, 2 for curve-fit instability and 2 for absence of stimulus-locked motion. 49 recordings from 20 participants were analysed (ADM n = 17, APB n = 18, TA n = 14); 20 of 22 participants contributed at least one valid recording.

Video was acquired at 127 +/- 15 frames per second (mean +/- SD), corresponding to a temporal resolution of 8.0 +/- 1.3 ms. Recordings contained a median of 105 stimuli (IQR 95-128) spanning a median 3.7-11.4 mA, delivered at a median of 77 distinct current levels (IQR 68-88) when recorded currents are grouped at a 0.02 mA tolerance.

### Activation thresholds

Examples of mechanical and electrical activation thresholds are shown per recording in Figure 3.

**Figure 3.**
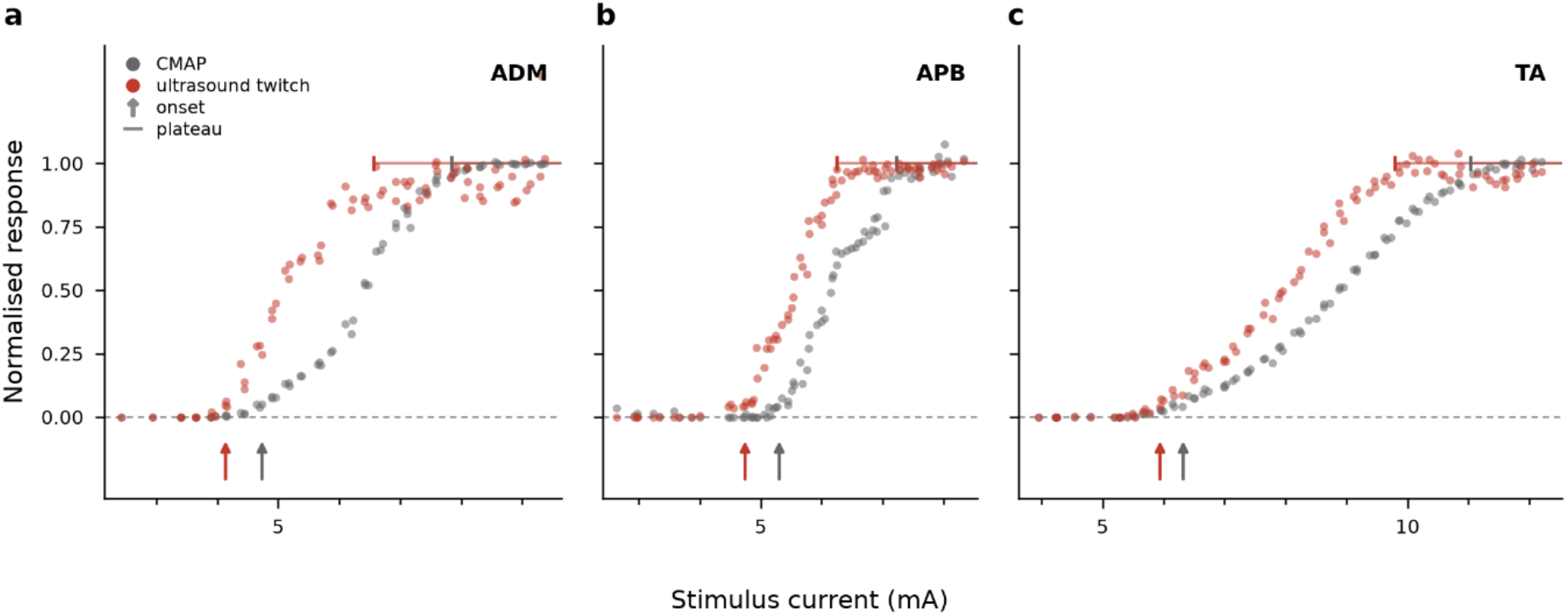
Example normalised CMAP and mechanical (ultrasound) twitch recruitment curves with increasing stimulus current. These examples show earlier threshold of muscle twitch prior to CMAP.

Threshold median values (CMAP/ultrasound) were 6.84/5.82 mA in ADM, 5.28/4.84 mA in APB, and 10.34/9.83 mA in TA. The within-recording difference (CMAP minus ultrasound; positive indicates a lower ultrasound threshold) was +0.27 mA in ADM (p = 0.009), +0.08 mA in APB (p = 0.001) and +0.21 mA in TA (p = 0.011). Adjusted across the six primary within-muscle tests, all three differences remained significant (ADM p = 0.013, APB p = 0.003, TA p = 0.013).

Expressed as a proportion of the motor unit recruitment range, the difference was 7.6% in ADM (p = 0.008), 2.9% in APB (p = 0.0008) and 5.1% in TA (p = 0.017).

Because the model-free analysis involves no curve fitting, it can also be computed for the recordings excluded for curve-fit instability. On that larger set of 51 recordings the same pattern holds: ADM +0.24 mA (n = 18, p = 0.048), APB +0.08 mA (n = 18, p = 0.001), TA +0.21 mA (n = 13, p = 0.011).

### Recruitment curve

The EC50 difference (CMAP minus ultrasound) was +1.07 mA in ADM (p = 0.002), +0.46 mA in APB (p = 0.003) and +0.90 mA in TA (p = 1.2 × 10^−4^). The per-recording EC50 difference correlated with the model-free threshold difference (Spearman rho = 0.71, p = 1.3 × 10^−8^, n = 49).

The plateau (the current at which the response reached 95% of its range) median values (CMAP/Ultrasound) were 9.80/7.78 mA in ADM, 7.81/7.47 mA in APB, and 15.86/13.65 mA in TA. The within-recording difference (CMAP minus ultrasound) was +0.99 mA in ADM (p = 0.020), +0.81 mA in APB (p = 0.008) and +1.21 mA in TA (p = 2.4 × 10^−4^). Adjusted across the same six tests, all three differences remained significant (ADM p = 0.020, APB p = 0.013, TA p = 0.0015).

### Motor unit twitch index

A motor unit twitch index was measurable in 32 of the 49 accepted recordings; the remaining 17 (11 participants) resolved fewer than five amplitude increments above their own noise floor. Values are shown per recording in Figure 4A.

**Figure 4.**
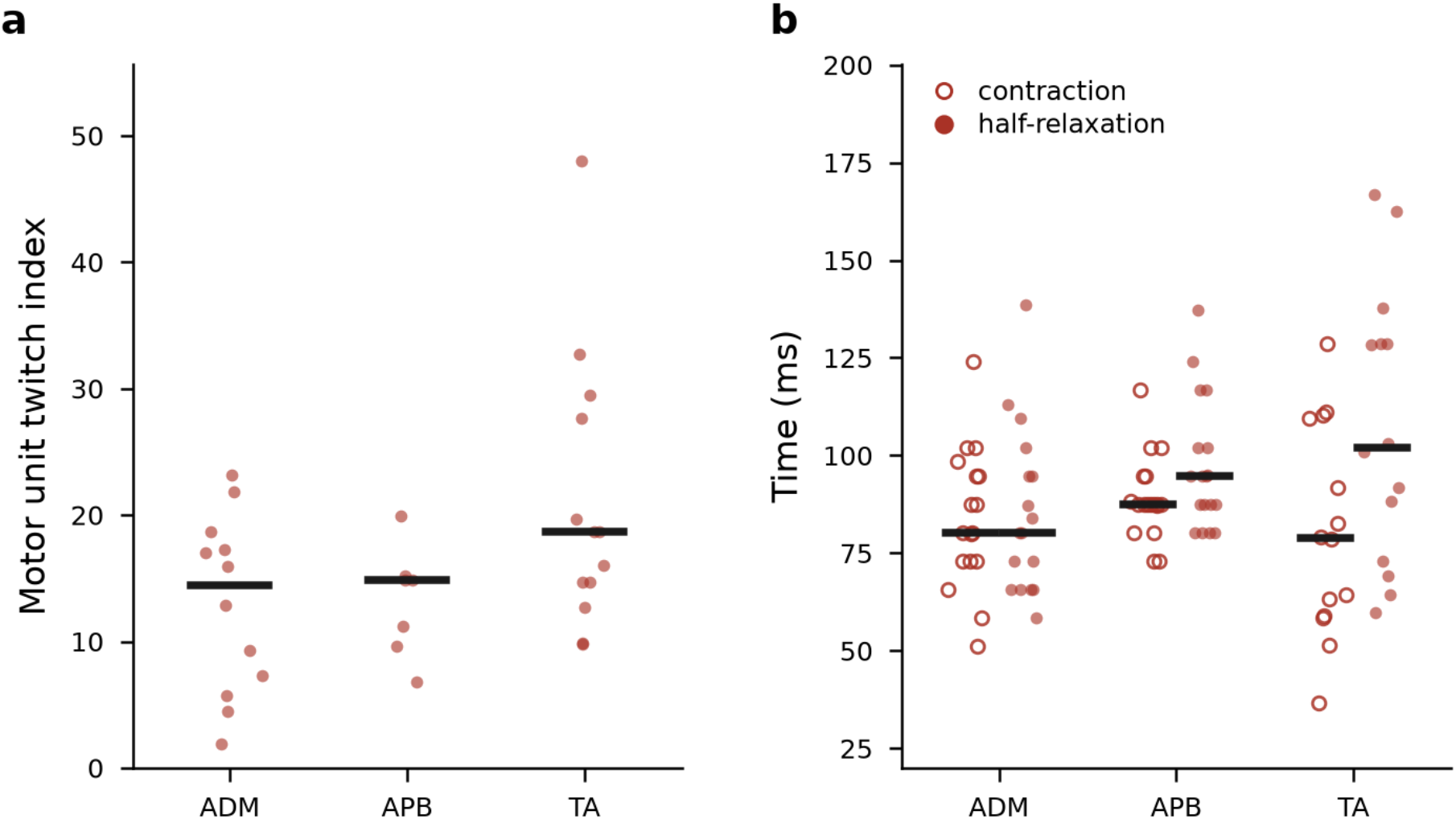
A-Motor unit twitch index values calculated using the primary model (Hill B) for each muscle. B-contraction and half-relaxation times per muscle. Line indicates median value.

The index was 14.4 in ADM (n = 12, IQR 6.9-17.6), 14.9 in APB (n = 7, IQR 10.4-15.0) and 18.6 in TA (n = 13, IQR 14.7-27.7).

TA had the highest median index under every alternative estimator examined (Hill/Gompertz/Weibull/probit × Rule A/B). However, the index correlated with the twitch signal-to-noise ratio across recordings (Spearman rho = 0.54, p = 0.001), which was highest in TA, so the between-muscle difference cannot be separated from a difference in signal quality. Full comparison of different estimators is shown in Supplementary Table 1.

Precision of the index at the level of a single recording is limited. The bootstrap interval (trials resampled within stimulus level) spanned a median of 40% of the point estimate (IQR 22-51%).

### Twitch timing

Contraction time and half-relaxation time are shown in Figure 4B. Median contraction time was 80 ms in ADM (IQR 73–95), 88 ms in APB (IQR 87–93) and 79 ms in TA (IQR 60–105). Median half-relaxation time was 80 ms in ADM (IQR 66–95), 95 ms in APB (IQR 88–103) and 103 ms in TA (IQR 77–129).

Timing was measured in a median of 36 trials per recording (IQR 21–48); within-recording trial-to-trial variability was 5.3 ms for contraction time and 8.3 ms for half-relaxation time.

## Discussion

We have shown that the twitch parameters of muscle can be measured directly using clinical ultrasound, and that twitch magnitudes recruit in parallel with CMAP amplitude.

The mechanical recruitment threshold was lower than the electrical threshold by a small margin (about 0.2 mA, or 3–8% of the electrical recruitment range) in all three muscles. A likely contributor is the differing fields of view of the two modalities. sEMG is spatially biased toward superficial motor units, with signal amplitude attenuating rapidly with depth. In contrast, the ultrasound field of view penetrates the full muscle depth, allowing for the detection of deep motor unit contractions that may be electrically silent or attenuated on the surface. Optical flow may therefore register the first recruited units, wherever they lie in the muscle cross-section, before a measurable surface potential appears. Furthermore, muscle twitch that occurs proximal or distal to the sampled region of muscle will still produce indirect contraction of the sampled tissue. Motor units have a limited longitudinal spatial extent^19^ so it is likely that some of the dissociation arises from longitudinal distance, and not just surface depth.

Conversely, the earlier plateau of the twitch signal compared to the CMAP, seen in all three muscles, likely reflects the physiological limits of mechanical temporal summation or the frame-rate constraints of the imaging system. Whilst electrical potentials can continue to summate linearly with additional recruitment of consecutive motor units, the mechanical twitch is limited by tissue compliance and the fusion of discrete motion events, which may result in an apparent saturation of the motion signal despite continued electrical recruitment.

The motor unit twitch index is the mechanical analogue of incremental MUNE and should not be read as an absolute motor unit count. Values of 14-19 are an order of magnitude below electrophysiological estimates for the same muscles (typically 100-250 by MUNIX^20^ or MScanFit^21^) and the shortfall is informative about what is being measured. The denominator is positively biased; only increments exceeding twice the noise floor are accepted, so the smallest twitches are excluded and the accepted increments are more likely to be composite of multiple units. Consistent with this, the index tracked signal-to-noise ratio across recordings, and the muscle with the cleanest signal returned the highest values. The numerator is biased downward by the nonlinear summation of twitch displacement which produces the earlier mechanical plateau.

The index is therefore best understood as a dynamic-range measure of mechanical recruitment, bounded by signal-to-noise ratio, and between-muscle differences cannot yet be attributed to differences in unit number. Approaching a true count would require a lower noise floor, correction for mechanical summation, or model-based fitting of the twitch recruitment scan.

It should be noted that whilst twitch time is derived from the point of zero-crossing of the twitch velocity, which therefore corresponds to maximal displacement as in traditional in vitro measures of twitch time, the half-relaxation time is measured from relaxation velocity, which may not be directly equivalent. With that said, the calculated twitch (79-88ms) and relaxation (80-103ms) times are similar to values produced in prior work measuring with force transducers directly on the muscle belly^22^, motor unit MRI of tibialis anterior^23^, and high-frame rate ultrasound^24^.

Motion was quantified with dense rather than sparse optical flow. Sparse methods track a set of salient (bright) features selected by a corner detector; in B-mode muscle images these lie preferentially on fascial planes and the bone surface, whose motion differs from that of the fascicles. These features may move in or out of plane of the ultrasound with deformation, so that tracked points drop out or jump. The number and location of surviving features then varies with gain, depth, echogenicity and muscle. Dense flow estimates displacement over the whole imaged cross-section, so the measurement domain is fixed by the field of view rather than by feature detection. It also makes our noise-suppression strategy possible: the spatial template and coherence score rely on genuine muscle motion being spatially coherent across the whole field while speckle-induced flow is not.

Several limitations must be acknowledged. The studied population was young and does not reflect muscle activity across the age spectrum. The analysis relies on 2D cross-sectional tracking, meaning out-of-plane dynamics such as longitudinal shortening, may be underestimated. Additionally, whilst the ∼127 FPS capture rate reflects standard clinical capabilities, it falls short of high-speed research sonography (>2000 FPS), limiting the precise resolution of rapid contractile properties. Furthermore, despite the advantages of dense flow tracking, it is also known to perform worst at abrupt discontinuities of motion and in textureless regions^25^, both of which may occur in extremes of muscle movement or disease states.

There are specific limitations relating to the motor unit number index measurement proposed. We could only produce a measurable index in 32/66 recordings overall due to the strict criteria we imposed for signal quality in order to overcome the further limitation that this value was dependent upon the signal to noise ratio seen.

Many clinical neurophysiology labs now have ultrasound devices in their department for assessing the structural properties of nerve & muscle, and newer EMG devices have integrated, or adjacent, neuromuscular ultrasound capabilities. Whilst this study has used research neurophysiology hardware to implement a custom utility with strict temporal alignment between ultrasound frames and electrical activation, a similar programme could be easily implemented with clinical devices at known frame rates, negating the need for temporal alignment.

Future studies must validate the performance of optical flow tracking in disease states characterised by altered muscle echogenicity, such as fibrosis or fatty infiltration seen in myopathic or neurogenic disease. In myopathy, the compound twitch parameters measured here were shown half a century ago, using force transducers on intrinsic hand muscles, to change in muscle disease: in Duchenne dystrophy the contraction disorder preceded any EMG abnormality in the same muscle, with prolonged half-relaxation^10,26^, whereas in myotonic dystrophy the contraction time was shortened^27^. Those methods required limb fixation and a transducer, and were never adopted clinically; the present method recovers the same parameters from the muscle belly with equipment already commonplace in neurophysiology departments. In neurogenic disease, unit number falls while the twitch force of surviving units changes with disease stage^28,29^, so both the index and the single motor unit twitch from which it is derived are expected to change.

Clinical ultrasound thus provides a non-invasive measure of the mechanical output of the motor unit population across the depth of the muscle, including its recruitment, twitch kinetics and departure from the electrical response. In doing so it opens the contractile properties of individual muscles to routine, repeatable assessment in health and disease.

## Data Availability

Anonymised data are available upon reasonable request

---

**Table 1.** Motor unit recruitment parameter (initial recruitment threshold, EC50 and plateau) differences between ultrasound and CMAP; motor unit twitch indices, and muscle twitch parameters for three muscles; Abductor digiti minimi (ADM), Abductor pollicis brevis (APB) and tibialis anterior (TA)

| Outcome | Unit | ADM | APB | TA |
| --- | --- | --- | --- | --- |
| Threshold gap (CMAP - US) | mA | +0.270<br>(p=0.009) | +0.079<br>(p=0.001) | +0.211<br>(p=0.011) |
| Threshold gap, relative | % of recruitment range | +7.6 | +2.9 | +5.1 |
| 4PL EC50 gap | mA | +1.07<br>(p=0.002) | +0.46<br>(p=0.003) | +0.90<br>(p=0.0001) |
| Plateau gap (CMAP - US) | mA | +0.988<br>(p=0.020) | +0.811<br>(p=0.008) | +1.208<br>(p=0.0002) |
| Plateau gap, relative | % of range | +14.5 | +11.1 | +12.5 |
| Motor unit twitch index (Hill B) | Index (IQR) | 14.4 (6.9–17.6) | 14.9 (10.4–15) | 18.6 (14.7–27.7) |
| Contraction time | ms | 80 | 88 | 79 |
| Half-relaxation time | ms | 80 | 95 | 103 |

**Supplementary Table 1.** Motor unit twitch index values using different statistical measures

| Model | Rule | Measurable /49 | ADM n | ADM median (IQR) | APB n | APB median (IQR) | TA n | TA median (IQR) |
| --- | --- | --- | --- | --- | --- | --- | --- | --- |
| Hill | A | 49 | 17 | 14.9 (9.0–18.7) | 18 | 16.6 (12.5–19.7) | 14 | 24.2 (20.0–28.0) |
| Gompertz | A | 49 | 17 | 14.6 (9.0–18.3) | 18 | 16.7 (13.2–19.6) | 14 | 24.8 (20.3–29.0) |
| Weibull | A | 49 | 17 | 13.7 (9.0–18.6) | 18 | 16.0 (13.1–18.8) | 14 | 22.1 (19.9–26.5) |
| Probit | A | 49 | 17 | 15.4 (9.0–18.8) | 18 | 16.3 (12.3–19.2) | 14 | 23.7 (19.8–28.3) |
| **Hill** | **B** | **32** | **12** | **14.4 (6.9–17.6)** | **7** | **14.9 (10.4–15.0)** | **13** | **18.6 (14.7–27.7)** |
| Gompertz | B | 18 | 7 | 17.7 (10.0–22.4) | 2 | 16.1 (15.9–16.3) | 9 | 26.4 (19.2–32.6) |
| Weibull | B | 38 | 14 | 11.6 (6.8–17.6) | 10 | 11.1 (7.8–13.7) | 14 | 15.7 (10.6–24.2) |
| Probit | B | 27 | 9 | 15.4 (6.8–19.0) | 6 | 12.3 (10.8–17.5) | 12 | 18.4 (13.9–27.2) |

## Notes

**Funding statement** This work was supported by an Accelerator Award from the Newcastle Neuroscience Fund. This work was funded by the NIHR Newcastle Biomedical Research Centre. The NIHR Newcastle Biomedical Research Centre is a partnership between Newcastle University and Newcastle Hospitals, funded by the National Institute for Health and Care Research (NIHR). This work presents independent research funded and supported by the NIHR Newcastle Biomedical Research Centre. The views expressed are those of the author(s) and not necessarily those of the NIHR or the Department of Health and Social Care.

### Competing Interest Statement

The authors have declared no competing interest.

### Author Declarations

Faculty of Medical Sciences Ethics Committee of Newcastle University gave ethical approval for this work.

